# Improving conversations about Parkinson’s dementia

**DOI:** 10.1101/2023.11.02.23297975

**Authors:** Ivelina Dobreva, Joanne Thomas, Anne Marr, Ruairiadh O’Connell, Moïse Roche, Naomi Hannaway, Charlotte Dore, Sian Rose, Ken Liu, Rohan Bhome, Sion Baldwin-Jones, Janet Roberts, Neil Archibald, Duncan Alston, Khaled Amar, Emma Edwards, Jennifer A. Foley, Victoria J. Haunton, Emily J. Henderson, Ashwani Jha, Fiona Lindop, Cathy Magee, Luke Massey, Eladia Ruiz-Mendoza, Biju Mohamed, Katherine Patterson, Bhanu Ramaswamy, Anette Schrag, Monty Silverdale, Aida Suárez-González, Indu Subramanian, Tom Foltynie, Caroline H. Williams-Gray, Alison J. Yarnall, Camille Carroll, Claire Bale, Cassandra Hugill, Rimona S. Weil

## Abstract

**Background:** People with Parkinson’s disease (PD) have an increased risk of dementia, yet patients and clinicians frequently avoid talking about it due to associated stigma, and the perception that “nothing can be done about it”. However, open conversations about PD dementia mean that people with the condition can access treatment and support, and are more likely to participate in research aimed at understanding PD dementia.

**Objectives:** To co-produce information resources for patients and healthcare professionals to improve conversations about PD dementia.

**Methods:** We worked with people with PD, engagement experts, artists, and a PD charity to open up these conversations. 34 participants (16 PD; 6 PD dementia; 1 Parkinsonism, 11 caregivers) attended creative workshops to examine fears about PD dementia and develop information resources. 25 PD experts contributed to the resources.

**Results:** While most people with PD (70%) and caregivers (81%) shared worries about cognitive changes at the workshops, only 38% and 30% respectively had raised these concerns with a healthcare professional. 91% of people with PD and 73% of caregivers agreed that PD clinicians should ask about cognitive changes routinely through direct questions and perform cognitive tests at clinic appointments. We used insights from the creative workshops, and input from a network of PD experts to co-develop two open-access resources: one for people with PD and their families, and one for healthcare professionals.

**Conclusion:** Using artistic and creative workshops, co-learning and striving for diverse voices, we co-produced relevant resources for a wider audience to improve conversations about PD dementia.

## Introduction

Although Parkinson’s disease (PD) is primarily a movement disorder, associated dementia is common, and affects nearly 50% of patients within 10 years of diagnosis [1]. Mild cognitive impairment, where cognitive deficits are present but do not impact day-to-day functioning, is present in 19-42% of newly diagnosed patients [2]. Dementia is diagnosed in PD when cognitive impairment becomes severe enough to affect the ability to carry out activities in everyday life. It is associated with poorer quality of life and higher levels of caregiver burden compared to PD without dementia [3], and double the level of financial burden compared to other dementias [4]. Despite the high prevalence and important practical implications of PD dementia, there is a lack of awareness of dementia symptoms in people with PD and their families and caregivers [5, 6]. In a survey of 209 carers, less than half (48%) were aware that people with PD are at increased risk of developing dementia [5]. Many people with PD and their caregivers have described feeling “left in the dark” due to lack of suitable information on dementia, and have described current memory services as disjointed, nonspecific and undersourced [7]. Of great concern, in a survey of 74 PD experts and healthcare professionals across the UK, only 14% said their training had prepared them well to provide high-quality care for people with PD-related dementia [5]. Cognitive function in PD was selected as a “community priority of major unmet need” by the PD Foundation Community Choice Research Award Program [8], and the lack of discussion about cognitive change was emphasised in a working group of experts convened in response to this [8]. Key barriers to conversations about dementia in the wider population are: 1) the stigma of a dementia diagnosis, including both the fear around the diagnosis, and worry about social exclusion; 2) the perception that treatment options are limited; and 3) that there is little benefit in early diagnosis or detection of dementia [9]. These factors are likely to be even greater in PD, where there is already a perceived stigma of the diagnosis of PD itself [10].

Barriers to help-seeking for cognitive problems are complex and multifaceted and are often considered to be more common in Black and other minoritized ethnic populations [11]. In a recent study, Black African and Caribbean community members described dementia as “a white person’s illness”, expressed views about the futility of seeking help for cognitive symptoms when there is no cure, and raised concerns about maintaining personal affairs private [12]. A review of the experiences of dementia in people of Black African and Caribbean backgrounds highlighted dissatisfaction due to inappropriate and perceived disrespectful treatment as an important barrier to help-seeking [13]. Studies in South Asian communities have also reported barriers to accessing services, including stigma associated with dementia, lack of culturally appropriate services, and preference for culturally aligned coping strategies [14]. To avoid stigma for the family, members from South Asian communities have been reported to provide care for affected family members privately rather than seek medical or social care assistance [14].

There are also challenges to discussing dementia in the clinic for healthcare professionals managing people with PD. A recent multidisciplinary symposium on unmet needs in cognitive health in PD found that cognition is not routinely or consistently assessed in the PD clinic, in contrast to motor function [8]. Barriers to clinicians initiating conversations about dementia in the PD clinic include a perception that there is nothing that can be done about PD dementia; lack of confidence in having conversations [5, 15]; lack of standardised ways to assess cognition and elicit cognitive symptoms in PD [8]; fear of inducing worry in people with PD or PD dementia [15]; and lack of time during appointments.

However, if dementia is not discussed in the PD clinic, people with PD dementia will not receive appropriate treatment or be able to access the support that is vital for them and their families. Receiving an earlier diagnosis of dementia can allow people with PD and their families to prepare for the future and to adequately make plans for care. Creating opportunities to discuss dementia-related issues can also help to engage patients in ongoing research and clinical trials in the PD dementia field.

To address the challenge of talking about dementia in PD, we worked together with people with PD and their families to 1) identify the roots and triggers of discomfort linked with talking about dementia; and 2) to find out how and when people wanted to hear about dementia in the clinic. We examined these issues during creative workshops, using artistic methods which offer the potential to mediate verbal delivery and generate multifaceted data. We took several steps to ensure that we included a diverse range of voices of lived experience, so that our findings would be more widely applicable to different groups. We used outputs from these workshops to form the basis of a pair of information resources to support dialogue and awareness of dementia in PD: one for people with PD and their families; and one for healthcare professionals to support these conversations in the clinic. We then held a series of focus groups and received feedback from people with lived experience and 25 PD experts to co-develop and refine these resources further. Both resources are now openly accessible as part of Parkinson’s UK’s information resources [16, 17]. We describe here the process of co-developing these information resources to ensure they would be widely accessible, well-targeted and practical.

## Methods

The project involved three stages:

1. Creative workshops with people with PD, PD dementia and their caregivers, to identify the roots and triggers of discomfort around conversations about dementia in PD, and to find out how, and when, people would like to receive information about PD dementia in the clinic. The outputs of these workshops formed the first draft of the information resources.
2. Focus groups with people with PD, PD dementia and their families, and with PD experts, to refine the information resources.
3. Wider consultation to access additional feedback and input into the resources from PD experts and the Parkinson’s community.

### Ethical approval and consent

The study was approved by the London Chelsea NHS Research Ethics Committee and participants provided written informed consent before participating. See also supplemental data for a GRIPP checklist considering patient public involvement for this study.

### Participants

We recruited participants between May and June 2022 from PD clinics at the National Hospital for Neurology and Neurosurgery, through Parkinson’s UK, and through previous study participants and community information events about research participation. We made efforts to ensure we included a diverse range of people with PD. To achieve this, we included a researcher on our core team (MR) whose work focuses on the experience of dementia and other chronic conditions in Black African, Caribbean, and other minoritized ethnic communities. MR guided our recruitment plans and materials and provided input throughout every stage of the project. Our goal for recruitment was at least 50% Black and other minoritized ethnic participants who did not identify as White. Inclusion criteria were a diagnosis of PD (or parkinsonism) and their carers/partners. We aimed to involve people with PD who were at different stages of progression: some with no symptoms of cognitive involvement, and others who already had cognitive symptoms or a PD dementia diagnosis. This allowed us to examine attitudes at different stages of the PD dementia journey. There were no exclusion criteria, but there were a few conditions which may have prevented participation (e.g. other medical or psychiatric conditions interfering with participation). We collected basic demographic information, including ethnicity and diagnosis.

### Stage 1: Creative workshops

We divided participants into two groups: predominantly PD without dementia (total *n*=14; comprising *n*=9 people with PD; and *n*=5 caregivers); and predominantly people with PD dementia (total *n*=20; comprising *n*=13 people with PD and *n*=7 caregivers). These groups aimed to allow participants to discuss attitudes towards dementia more freely with others at a similar disease stage. However, we were limited by participant availability on particular workshop days, so each group had some people with PD and PD dementia. Each group attended two workshops over two months, which were held at Central Saint Martins School of Art (part of the University of the Arts London). The workshops were designed by the core team which included artists experienced in using creative methods to facilitate conversation; a person with lived experience of PD; PD clinicians; and public engagement experts. Each workshop lasted approximately 120 minutes, and included a gentle physical warm-up led by a Pilates instructor, an ice-breaker art exercise, the core creative session (45min) and a “show and tell” discussion facilitated by two artists in the core team (45min).

During all workshops, clinicians, researchers, and public engagement experts also participated with the creative elements. On-site breakout-rooms and nominated staff members were available if participants felt upset, distressed, or simply wanted to take a break during dementia discussions and needed to step away from the session, but these were not required. Helpline and support information from Parkinson’s UK was also shared during and after workshops.

Workshop 1 explored beliefs, fears and attitudes people with PD and their family members held about dementia. Using paint, drawing and collage, and supported by artists, participants shared their associations with PD dementia. Through these media, they were encouraged to explore their attitudes towards receiving a diagnosis and consider their perceptions about living with PD dementia. This included any difficulties talking to others about the diagnosis. Participants were invited to create as many artistic outputs as they wished, which included paper cut-outs as well as paintings. After the creative session, participants provided descriptions of their artwork in a “show and tell” discussion.

In Workshop 2, participants were invited to create artworks using paint and collage to share how they would like to learn about dementia from healthcare professionals. Key questions they were asked to consider were: *“How would you prefer to learn about dementia?”*; “*At what stage?”; “From whom?”;* “*Where*?”; “*In what format*?”; and “*What does excellent PD dementia support look like?”*. As in the first workshop, participants were encouraged to create as many outputs as they wished and provided descriptions of their artwork.

During the “show and tell” session of each workshop, members of the core team documented comments and art descriptions made by study participants. See Fig. 1 for creative outputs from the workshops. A thematic analysis of the notes and artwork was performed, and the emerging themes were used to guide the development of a pair of information resources: one for people with PD and their families and one for healthcare professionals.

**Figure 1.** Outputs from the creative workshops that informed the content of information resources.

### Feedback Surveys

Participants completed an anonymised questionnaire of open and closed-ended questions exploring attitudes and fears regarding PD dementia before each workshop. We additionally collected anonymised questionnaire responses about the experiences during the project, and change in attitudes to PD dementia as a result of taking part in the project (see Supplemental material for questionnaires). A thematic analysis of the free text was undertaken as well as quantitative analysis of available data.

### Stage 2: Focus groups

We then held a series of focus groups with the workshop participants, to refine and review the information resources, which had been developed using the creative outputs and discussions during the workshops. Participants remained in the same groups they attended the workshops in, namely PD without dementia (total *n*=6; comprising *n*=6 people with PD); and people with PD dementia and their caregivers (total *n*=8; comprising *n*=2 carers and *n*=6 people with PD dementia). The focus groups were divided into three sessions. The first focus group session sought feedback on content of the information resources. The second and third sessions sought feedback on overall appearance, design, and structure of the information resources.

In addition, we held two focus groups with a total of 12 healthcare professionals (comprising 2 PD nurses, 1 psychologist, 3 geriatricians, and 6 neurologists). These were recruited from PD professional networks. The purpose of these focus groups was to review and co-develop each of the information resources, and to ensure that information in the resources reflected current best practice. Specifically, for the patient-focused information resource, we sought feedback on the communication of prevalence and risk factors for PD dementia, inclusion of advice about exercise, feedback on the appearance of the resource, and how it might be used in the clinical setting. For the healthcare professional resource, we sought feedback on cognitive assessment, diagnostic procedure and treatment strategies, and overall tone of the resource.

### Stage 3: Wider Consultation

A final stage was to test the near-final draft booklets with people who had not been involved in the project to date, to ensure that the content was widely accessible, complete, and relevant. We held a focus group with people with PD (*n* =8), recruited through Parkinson’s UK, who had not attended the workshops. Participants were provided with the patient-facing resource prior to the focus group to give them a chance to consider it fully. At the focus group, they were asked for their feedback on the resource content, utility, use of language, format and layout.

We sought the opinions of an additional 13 PD healthcare professionals to review the information resources, to check the factual content, and advice, and consider the way information was communicated. We included eight neurologists, one psychologist, two physiotherapists, an occupational therapist, and one psychiatrist in this feedback stage.

To measure impact of the resources, we collected data on number of online downloads, and number of requests for printed versions, in the three months following their release.

## Results

34 people affected by PD took part in the four creative workshops. These included 16 people with PD, diagnosed according to UK Brain Bank criteria, 6 with PD dementia (diagnosed clinically [18], 1 person with parkinsonism, 11 family members or caregivers (9 spouses and 2 siblings). Mean disease duration was 6.76 years, SD=3.96. None of the people with PD had a formal diagnosis of PD-MCI. We included a person with parkinsonism who was still going through a diagnostic process, as this reflects the experience of people with PD, who may receive a definitive diagnosis at a later stage. 43% of people with PD and 46% of caregivers self-identified as Asian, Black, or Mixed ethnic group. In addition, 2 clinicians, 4 researchers, 4 public engagement experts, and 4 art facilitators participated in the workshops. Characteristics of participants affected by PD are presented in **Table 1**.

**Table 1.** Participants demographics for the workshops.

| **Attribute** | **People with**<br>**Parkinson's *n* = 23** | **Caregivers *n* = 11** |
| --- | --- | --- |
| Age, years | 69.1 (5.6) | 69.5 (6.2) |
| Male, n (%) | 14 (60.68%) | 0 |
| Female, n (%) | 9 (39.13%) | 11 (100%) |
| Ethnicity, n (%) |  |  |
| Asian | 7 (33.3%) | 1 (9%) |
| Black | 2 (9.52%) | 3 (27.27%) |
| Mixed | 0 | 1 (9%) |
| White | 12 (57.14%) | 6 (54.54%) |
| Not disclosed | 2 (9.52%) | 0 |
| Time since Parkinson's diagnosis, years | 6.76 (3.96) | N/A |

Age and time since diagnosis variables shown in mean (SD). Other data reported in number of participants, and percentage.

### Themes emerging from the workshops

Two main themes were identified from creative outputs and discussions in workshop 1: (1) fear of PD dementia symptoms; (2) the stigma associated with receiving a diagnosis, including a worry of exclusion from communities. Words linked with dementia included “scared” and “struggle”. Participants expressed concerns over loss, specifically loss of identity, control, independence, and social life. People also described fear of not being seen as an individual, but as a “person with a disease”.

Stigma emerged as tightly linked to fear, with participants describing their reluctance to share receiving the diagnosis with friends or distant family. In particular, they expressed a worry about how they would be perceived in their community. This was especially pronounced for people from Black African and Asian communities.

Workshop 2 explored how participants would like to receive information about PD dementia. Participants described that they would like to receive this information from a medical professional, in a written format that is easily accessible, early in the disease journey. They also expressed the need for clear information on medication and how it can help in the context of PD dementia, as well as guidance on what support is available to them.

### Survey results on attitudes to dementia

Prior to the workshops, 70% of people with PD and 81% of caregivers shared that they had been worried about PD dementia. However, only 38% of people with PD and 30% of caregivers had previously raised these concerns with a healthcare professional. Reasons for not previously raising these worries were lack of opportunity, and fear of discussing the issue in front of their loved one (See Box 1).

#### Box 1.

**Descriptions of barriers to talking about PD dementia**

“I did not like to raise it [with a doctor] in front of my partner” (Caregiver)

“I volunteer with a wider PD group and while everything else is discussed, dementia isn’t. We are a very social group, and it would be very helpful to have a new way of talking about dementia without scaring everyone.” (Caregiver)

“I felt my consultant did not want to address the issue of dementia” (Participant with PD dementia)

[We had a] “good discussion, but short – only 10 minutes” (Participant with PD dementia)

“Fear – I am a Christian and I believe that what you talk about is what you get. I believe in God protecting my husband [from dementia]” (Caregiver)

Experiences of discussing dementia with healthcare professionals varied from “constructive”, to “not very helpful”. Caregivers found the conversation “quite easy but concerning”, “distressing, although expected diagnosis of dementia”, and “very helpful”. When asked if there was anything about how the PD dementia diagnosis could have been improved, one person with PDD and one caregiver wanted more information on the condition, one person with PDD did not wish to receive a diagnosis; and one felt their consultant was reluctant to discuss it. Three people with PDD responded that nothing could be improved.

90% of people with PD and 73% of caregivers agreed that clinicians should ask people with PD about thinking and memory changes routinely. People with PD and caregivers indicated that this should be done through direct and focused questions, routine memory tests, and more regular appointments to track changes (see Box 2).

#### Box 2.

**How people with PD want to talk about PD dementia**

“I prefer a direct question with a preamble by the clinician of how it is routine.” (Participant with PD)

“Point out what it can be like [when cognitive symptoms occur] to enable us to identify the issues early on.” (Participant with PD)

“Important to allow sufficient time to fully discuss the issue” (Participant with PD)

“Ask routinely and by explaining what symptoms may occur to bring awareness” (Participant with PD)

“Reviews need to be more regular, as we currently see our Parkinson’s nurse [only] once per year”. (Participant with PD)

“Sensitively, perhaps together with caregivers, as they would notice some early changes” (Participant with PD)

“Provide more explanations about possible symptoms and what further treatments or help there is” (Caregiver)

Two people highlighted the need for more time during consultation appointments, and the importance of caregivers in recognition of early changes. When asked if there are different words that could be used instead of dementia, most people with PD did not mind the use of the word “dementia” in facilitating conversations around thinking and memory.

### Survey results on experience of participating in the creative workshops

Feedback from people with PD and caregivers on participating in the creative workshops was overwhelmingly positive. They used the words “*emotional”, “enlightening”, “heart-warming”, “supportive”*, and *“brilliant”* to describe how it felt to take part (see Box 3). They described it as reassuring to hear other people’s similar experiences and learn from them. 80% of people shared that participating in the creative workshops had changed the way they view conversations on PD dementia. Participants described that they are now more ‘open to’ and ‘confident’ in holding these conversations and would find it easier to talk about this with their family or clinicians. All participants (100%) responded that they would take part in a similar project again, or would recommend taking part in a similar project to someone they know.

#### Box 3.

**Feedback on taking part in creative workshops to improve PD dementia conversations**

“Prior to [the] workshop I couldn’t quite understand how art could be used to discuss dementia - I was quite inspired and enlightened.”

“It has been eye opening to see that people are all so different [in the way they experience PD] and see things so differently and yet so coincidentally.”

“Being part of this project helped me understand that I am not alone in living with PD – others are going through the same experiences, and we can learn from each other.”

“Joining in the workshops brought a feeling that I was contributing to something that would be of real, tangible benefit to many.”

“I have learnt not to be afraid of talking about my feelings of dementia. Meeting others with similar experience definitely helps in making it easier to convey.”

### Content co-development for the information resources

We used the outputs from the workshops, as well as the surveys to form the basis of the two information resources: one for people with PD and their families, and one for healthcare professionals in specialist PD clinics.

We structured the content of the patient information resource into nine sections, beginning with what thinking and memory changes are like in PD, and why they happen. We included a section on how to raise worries about dementia with a specialist; information on medications; and details of support for caregivers, including advanced stages and end of life. The healthcare professional resource was designed to be of practical use in a clinic where time may be limited, with information on recognising and diagnosing PD dementia, as well as practical management guidance (see Fig. 2 for cover images of the information resources).

**Figure 2.** Front covers of the information resources for (A) people with PD; and (B) for healthcare professionals.

Both information resources have been adopted as resources on PD dementia by Parkinson’s UK, and are freely available to download from the Parkinson’s UK website [14, 15]. (Links available at the end of this manuscript). Within 3 months of their launch, the pages with these resources have already had 17,869 page views (13^th^ most viewed page on Parkinson’s UK’s site in that time period). Print orders for the patient booklet is 769, and 334 for the healthcare professional resource.

## Discussion

In this collaborative project, we aimed to identify roots and triggers of discomfort linked with dementia within the PD community and to co-produce a pair of information resources, for people with PD and healthcare professionals, to improve conversations about PD dementia. We found that although 70% of patients and 81% of caregivers had been worried about PD dementia, only 38% and 30% respectively had raised these concerns with a healthcare professional. Conversely, we found that during the workshops, people affected by PD were willing to talk and learn about dementia, and were open to discussing the risk of dementia and support available to them. Even though people with PD acknowledged that talking about dementia can be challenging, they recognised the benefits of doing so and thought that PD clinicians should routinely and directly ask patients about potential cognitive changes and use cognitive tests at regular clinic appointments.

Discussing dementia in a PD clinic where time is often limited, can be challenging. Conversations may be hindered by barriers for both people with PD and healthcare professionals. We found that using art and creative approaches helped people speak more freely and broach conversations they might have found difficult to bring up in other settings. Whilst this would not usually be practical for clinical settings, it allowed us to explore these issues and develop useful information resources. These will mean that people with PD dementia can access information on risk, timely diagnosis, treatment, and patient-centred support.

Working with artists and using a creative process for the workshop stage of this project enabled people with PD to feel comfortable and encouraged participants to open up in a way they may have found challenging in more conventional settings. Being able to deliver the workshops away from the hospital, together with clinicians and researchers, removed the usual patient-doctor hierarchies, and ensured that clinicians and researchers could become equal partners in this project, to address the challenge of how to talk about PD dementia.

There has been an increased recognition of the utility of arts-based interventions (including art-making and dance) for people living with dementia and their caregivers. Art-making has been linked to an increase in wellbeing, improvement in social participation and improvement in psychological health [19, 20]. Creative expression and artistic activities such as painting have also been found to be an important way for people with dementia to express and access emotions even in the presence of cognitive decline[21, 22]. In PD, art-based interventions have shown improvements in motivation and creativity [23], and participation in dance programmes leads to improvement in motor symptoms and wellbeing [24, 25]. To our knowledge, this is the first project to apply creative processes to facilitate conversations about PD dementia. However, similar creativity-based methods have been used to explore perceptions of clinical trials in ethnic minority populations [26], and to understand South African women’s perspectives on reproductive health [27]. Both of these projects demonstrated the benefits of creative approaches to facilitate challenging conversations.

Using the workshops’ outputs, we created two information resources, which benefited from input from experts in PD from a range of clinical settings around the UK. This meant we had the most up to date guidance on diagnosing and managing PD dementia, from experts across a range of disciplines. By disseminating these tools through Parkinson’s UK (the largest UK charity for PD), we can ensure they will be freely available, widely accessible and viewed as trustworthy and credible sources of information. It will also provide a framework for updates as new guidance and information becomes available for PD dementia.

## Strengths and limitations

Particular strengths of this project are that the voices of people with PD were at the core of all stages; and that we included a diverse range of people with PD, particularly those from ethnic minority backgrounds, who are often underrepresented in PD research. This meant that our resources could resonate with more people with PD, be useful to a wider audience, and that we could address issues, such as the stigma of PD dementia, that might not be raised with a more homogeneous group.

Although we aimed for diversity on a range of different aspects, this project was carried out in the UK. Factors such as cultural views on dementia, access to healthcare, and support infrastructure will vary in other parts of the world. Our group was also relatively young for PDD, with relatively short disease duration, so may not be fully representative of populations with PDD. Our workshops were intentionally small, to allow people to feel comfortable talking about dementia in a friendly environment. However, this meant that our sample size was relatively small, in common with qualitative research in general. Future work could apply similar approaches with a more global outreach, to develop more international guidance.

## Data Availability

All data produced in the present study are available upon reasonable request to the authors.

## Summary

Through a process of artistic workshops, co-learning and bringing in diverse voices, we co-developed two information resources to improve conversations about PD dementia: one for people living with PD, and one for healthcare professionals, that are freely available to be used by people with PD and healthcare professionals. We hope that these resources will help facilitate these challenging conversations and result in helping people living with PD dementia to access more timely diagnosis, treatment, and patient-centred support.

## Open access links to Parkinson’s dementia resources

Patient and family resource:

https://www.parkinsons.org.uk/sites/default/files/2023-07/CS3923%20Thinking%20and%20memory%20changes%20in%20Parkinson%27s_Lived%20expert%20toolkit_A5_Final_WEB.pdf

Healthcare professional resource:

https://www.parkinsons.org.uk/sites/default/files/2023-07/CS3923%20Detecting%20and%20managing%20Parkinson%27s%20dementia_Healthcare%20professional%20toolkit_Final_WEB_0.pdf

## Data Sharing

The data supporting the findings of this study are available from the corresponding author, upon reasonable request.

## Acknowledgments

We thank people with PD and caregivers for their time and invaluable feedback during all stages of this project; and to all PD specialists who fed back on both resources.

## Authors Roles

RW, CH, AM, RO, JR, JT and CB conceived and designed the project. MR consulted on recruitment plans, and ensuring participant representation from ethnic minority groups. ID recruited participants, analysed data and wrote the first draft of the manuscript. AM, RO, RW, JT, ID, MR, CD, KL, SR delivered the artistic workshops. JT, RW, ID, CD, MR, SBJ delivered the focus groups and wider consultation sessions. All authors wrote and approved the final version of the manuscript.

## Financial Disclosures of all authors (for the preceding 12m)

ID, JT, RO, MR, NH, CD, SR, KL, RB, SBJ, JR, NA, DA, KA, AJ, FL, CM, LM, ERM, BM, KP, BR, AS, MS, ASG, IS, TF, CC, CB, CH, report no disclosures for the preceding 12 months.

AM has received workshop consultancy honoraria from Crossover Labs

EE reports Honorariums from Bial and Neurology academy.

JAF has received research funding from Parkinson’s UK.

VJH has received travel support from Bial and research funding from Parkinsons UK and National Institute for Health and Care Research

EJH has received honoraria and / or travel support / advisory board contribution from Kyowa Kirin; Simbec Orion, Abbvie; Ever; Bial; and the Neurology Academy; and research funding from The Gatsby Foundation, Royal Osteoporosis Society, National Institute of Health Research and Parkinson’s UK.

CWG has received grant support from Cure Parkinson’s, Parkinson’s UK, the Rosetrees Trust, and consultancy fees from Evidera.

AJY has received honoraria from GE Healthcare, plus grants from Cure Parkinson’s, Lewy Body Society, EU IMI, Parkinson’s UK, Dunhill Medical Trust, NIHR

RSW has received speaking honoraria from GE Healthcare and Bial, and has provided consultancy to Therakind.

