## Supplementary Material for "Improving conversations about Parkinson’s dementia"

Supplemental Material

| Section and topic | Item | Reported on page No |
| --- | --- | --- |
| 1: Aim | Report the aim of PPI in the study | 6 |
| 2: Methods | Provide a clear description of the methods used for PPI in the study | 7-10 |
| 3: Study results | Outcomes—Report the results of PPI in the study, including both positive and negative outcomes | 10-14 |
| 4: Discussion and conclusions | Outcomes—Comment on the extent to which PPI influenced the study overall. Describe positive and negative effects | 15-16 |
| 5: Reflections/critical perspective | Comment critically on the study, reflecting on the things that went well and those that did not, so others can learn from this experience | 17 |

GRIPP2 short form

PPI=patient and public involvement

**Pre-workshop survey for people with Parkinson’s**

1. A. Have you been diagnosed with Parkinson's disease? Please circle one option:

Yes / No

B. If so, when were you diagnosed (month,year)?

2. Have you been worried about changes in thinking and memory? Please circle one option:

Yes / No / Don’t know

If so, in what way?

2. A. Have you raised this with your doctor? Please circle one option:

Yes / No / Don’t know

B. If not, why not?

C. If yes, how did you find this conversation?

4. Have you been diagnosed with Mild Cognitive Impairment or Parkinson's dementia? Please circle one option:

Yes / No / Don’t know

A. If so, when were you diagnosed (month, year)?

5. Is there anything about how the diagnosis was made that could have been done better?

6. A. Should doctors ask people with PD about thinking and memory changes routinely? Please circle one option:

Yes / No / Don’t know

B. If not, why not?

C. If yes, how should they do this?

7. Are there different words that doctors could use instead of "dementia" to help facilitate these conversations? What might these words be?

8. Why are you interested in participating in this project?

**Pre-workshop survey for carers and family members**

1. Are you the friend or family of a person who has been diagnosed with Parkinson's disease?

What is the nature of your relationship? (e.g. partner, child, friend..)

2. When were they diagnosed with Parkinson’s disease? (month, year)

3. Have you been worried about changes in their thinking and memory?

A. If so, in what way?

4.Have you raised this with their doctor? Please circle one option:

Yes / No / Don’t know

A. If not, why not?

B. If yes, how did you find the conversation?

5. Has your friend/family member been diagnosed with Mild Cognitive Impairment or Parkinson’s dementia? Please circle one option:

Yes / No / Don’t know

A. If so, when were they diagnosed (month,year)?

6. Is there anything about how the diagnosis was made that could have been done better?

7. Should doctors ask people with Parkinson’s about thinking and memory changes routinely? Please circle one option:

Yes / No / Don’t know

A. If not, why not?

B. If yes, how should they do this?

8. Are there different words that doctors could use instead of "dementia" to help facilitate these conversations? What might these word be?

9. Why are you interested in participating in this project?

**Feedback Survey**

1.
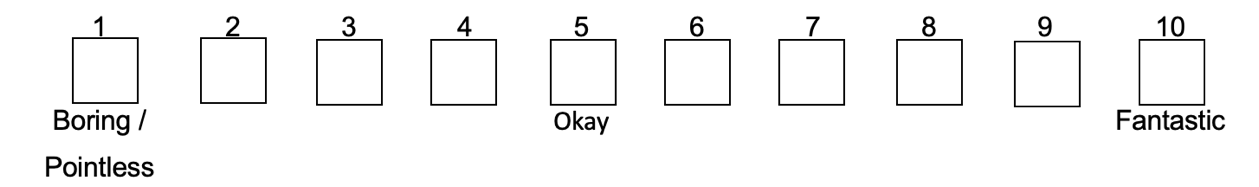
Overall, how was your experience taking part in the project?
A. How would you describe your experience in this project?
2.
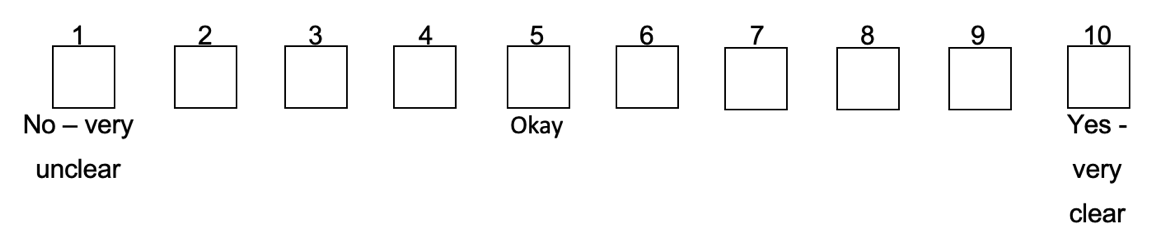
Where the aims of the session explained clearly? Tick one box.
3.
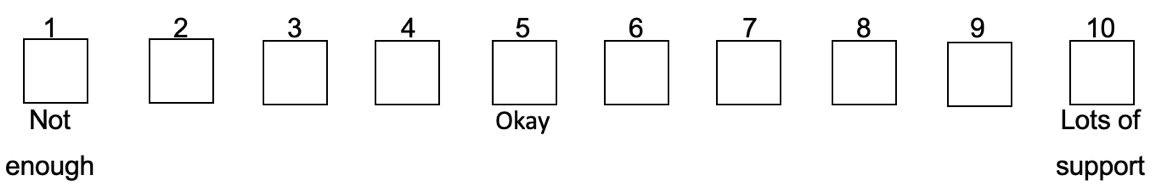
How supported did you feel by the organisers during the session? Please tick one box.
4.
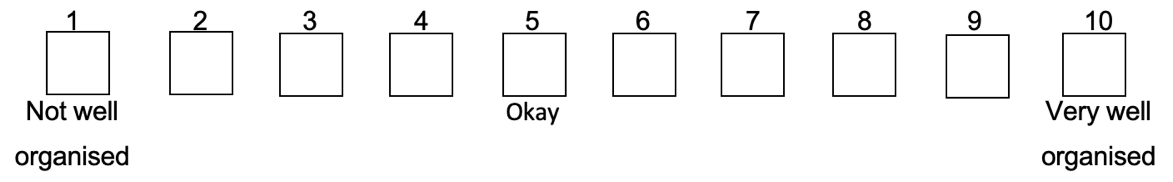
Do you feel the sessions were well-organised? Please tick one box.
5. What was good about being involved in the project?
6. What could have been better?
7. What do you feel you learned and/or gained from the project?
8. A. Since taking part, have you changed the way you view conversations about thinking and memory in Parkinson’s? Please circle one option

Yes / No	/	Maybe

B. Can you tell us a bit more about your answer above?
9.
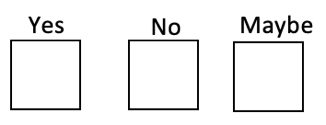
Would you be interested in taking part in a similar project again? Please tick one box.
10.
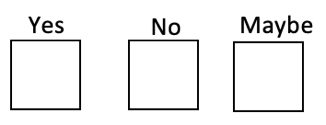
A. Would you recommend taking part in a project like this to someone you know? Please tick one box.
C. Why/Why not?

11. Do you have any other feedback or comments about your experience in the project?
